# Correcting prevalence estimation for biased sampling with testing errors

**DOI:** 10.1101/2021.11.12.21266254

**Authors:** Lili Zhou, Daniel Andrés Díaz-Pachón, Chen Zhao, J. Sunil Rao, Ola Hössjer

## Abstract

Sampling for prevalence estimation of infection is subject to bias by both over-sampling of symptomatic individuals and error-prone tests. This results in naïve estimators of prevalence (i.e., proportion of observed infected individuals in the sample) that can be very far from the true proportion of infected. In this work, we present a method of prevalence estimation that reduces both the effect of bias due to testing errors and oversampling of symptomatic individuals, eliminating it altogether in some scenarios. Moreover, this procedure considers stratified errors in which tests have different error rate profiles for symptomatic and asymptomatic individuals. This results in easily implementable algorithms, for which code is provided, that produce better prevalence estimates than other methods (in terms of reducing and/or removing bias), as demonstrated by formal results, simulations, and on COVID-19 data from the Israeli Ministry of Health.

## 1 INTRODUCTION

Estimation of disease prevalence is challenging. First, except for the hypothetical case of random errors, imperfect testing almost always distorts actual proportions. Second, it is not uncommon to have to derive estimates from samples that under-represent or fail to capture subpopulations that are at greatest risk or of interest. An example is estimating the general population prevalence of chronic hepatitis C (HCV) because of the challenges of sampling from subpopulations of former and current injecting drug users, the homeless or incarcerated. ^1^ Other examples include the over-representation of symptomatic individuals in a sample since these individuals are more likely to get tested than asymptomatic ones, with which the final estimates of prevalence inflates, since symptomatic individuals are also more likely to be truly infected than asymptomatic ones. ^2^

This situation became clear during the recent COVID-19 pandemic: besides usual discussions of the error rates of PCR and rapid tests, surveillance mechanisms have usually relied on convenience sampling or contact tracing. Therefore sampling bias was also present. In the case of convenience sampling, because it passively waits for symptomatic individuals to get tested, whereas asymptomatic individuals have few reasons to do so. As for contact tracing, because it actively pursues infected individuals, ignoring the non-infected almost altogether. Besides this, contact tracing has also raised questions on privacy and individual liberties. ^3,4,5^ Though this example corresponds to a non-probability COVID-19 sampling setting, the problem is of course more general. It applies not only to every form of prevalence estimation performed through testing —probabilistic or not— and even more general forms of selection bias. ^6,7,8,9^

Recently, Díaz-Pachón and Rao introduced a correction for oversampling of the symptomatic group. ^10^ It was a three-step procedure based on the assumption that all symptomatic individuals in the population were sampled and infected but it did not address the issue of imperfect testing (i.e. the presence of false positives and false negatives). This implies that the symptomatic and infected individuals in the sample corresponded to the total number of symptomatic individuals in the population. Thus the asymptomatic group in the population was the complement of the total of symptomatic individuals in the sample. The prevalence among the asymptomatic group was then obtained as a uniform random variable among the asymptomatic individuals in the population, with no resource to the sample.

In this paper a method that is stronger in all aspects is presented. First, it does not assume that *all* symptomatic individuals are sampled, only that symptomatic individuals are overrepresented in the sample. Second, sample values among the asymptomatic are used to produce an estimator of prevalence that is informed by evidence. Third, testing errors are considered. And fourth, the proposed correction is extended to stratified errors by symptom status.

The article can be summarized as follows:

1. The researcher observes, from a sample of size *N*_*T*_, the proportion of individuals whose test was positive. This constitutes the naïve estimator of prevalence 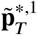.
2. The naïve estimator 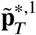 is biased in two ways. First, it is subject to testing errors; and second, there is sample bias because symptomatic individuals are more likely to be tested than asymptomatic ones.
3. Under the assumption that, from a different and independent sample, testing error rates are estimated for group *s* as 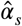 and 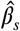 for false positives and false negatives, respectively, it is possible to obtain another estimator 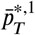 that corrects the effect of errors.
4. From 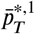 it is possible to obtain another estimator 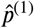 that reduces (and possibly removes) the sampling bias. This is achieved through applying the principle of maximum entropy to the fraction of symptomatic individuals with the disease, using the knowledge that symptomatic individuals are more likely to get tested.

With this summary and the information of Table 1, without having to go through the details that led to their derivation, the reader can obtain corrections following the steps of Algorithm 1 when no testing errors are present and all symptomatic individuals are sampled, Algorithm 2 when no testing errors are present and not all the symptomatic individuals are sampled, Algorithm 3 when testing errors are present and all symptomatic individuals are sampled (provided that the testing errors are estimated unbiasedly), and Algorithm 4 when there are testing errors and not all symptomatic individuals are sampled. Section 5 presents an example with real data. Proofs of all the results are consigned to the Appendix, as well as a set of simulations that assess the behavior of the four algorithms.

**TABLE 1.** A population of known size *N* is divided into symptomatic individuals (*s* = 1) and asymptomatic ones (*s* = 0), and non-infected individuals (*i* = 0) and infected ones (*i* = 1). The second column gives the notation for symptoms *s* and infection *i*, while the third column marginalizes symptoms, and the fourth one marginalizes infection. To facilitate reading, the notation is arranged as follows: **(a)** the letter *q* is only used for sampling probabilities; **(b)** all estimators are capped by tildes, bars, or hats; **(c)** bold caps refer to naïve estimators; **(d)** estimators not in bold are corrections of naïve estimators; **(e)** population proportions do not have any cap; **(f)** prevalence values are obtained replacing *i* in the last column by 1.

| Quantity | Symptoms $s$ and infection $i$ | Symptoms $s$ | Infection $i$ |
| --- | --- | --- | --- |
| | $I_s^{(i)}$ | $I_s$ | $I^{(i)}$ |
| population totals | $N_s^{(i)}$ | $N_s$ | $N^{(i)}$ |
| population proportion | $p_s^{(i)}$ | $p_s$ | $p^{(i)}$ |
| sampling probability | $q(I_s^{(i)})$ | $q(I_s)$ | $q(I^{(i)})$ |
| sampling probability approximation | $q^*(I_s^{(i)})$ | $q^*(I_s)$ | $q^*(I^{(i)})$ |
| sampling totals | $N_T^{s,i}$ | $N_T^{s,*}$ | $N_T^{*,i}$ |
| naïve estimator<br>(with errors and sampling bias) | $\tilde{\mathbf{p}}_T^{s,i}$ | $\tilde{\mathbf{p}}_T^{s,*}$ | $\tilde{\mathbf{p}}_T^{*,i}$ |
| naïve estimator<br>(only with sampling bias) | $\hat{\mathbf{p}}_T^{s,i}$ | $\hat{\mathbf{p}}_T^{s,*}$ | $\hat{\mathbf{p}}_T^{*,i}$ |
| corrected estimator for errors | $\bar{p}_T^{s,i}$ | $\bar{p}_T^{s,*}$ | $\bar{p}_T^{*,i}$ |
| corrected estimator for errors and<br>sampling bias | $\hat{p}_s^{(i)}$ | $\hat{p}_s$ | $\hat{p}^{(i)}$ |

## 2 SETTING

Consider a population 𝒫 of size *N* that is divided into four categories: asymptomatic and non-infected individuals, 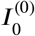, with size 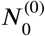; asymptomatic and infected individuals, 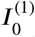, with size 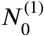; symptomatic and infected individuals, 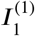, with size 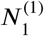; and symptomatic and non-infected individuals, 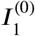, with size 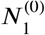. The population total *N* is known, whereas 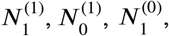, and 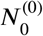 are unknown, though their sum is *N*.

The group of individuals with symptoms *s* in the population will be denoted by 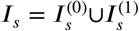, and its total by 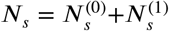, for *s* = 0, 1. Analogously, the group of individuals with infection status *i* in the population will be denoted by 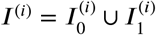 and its total by 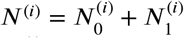, for *i* = 0, 1.

Now, 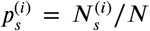 will be the proportion of individuals in the population with symptoms *s* and infection status *i*. More formally, define a random element 𝒮^*^ taking values in the set 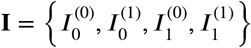, with density given by

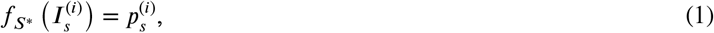

and 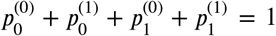. The proportion of individuals in the group *I*_*s*_ is then given by 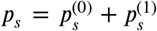, for *s* = 0, 1. And t^0^he pro^0^portion^1^ of in^1^dividuals in the group *I*^(*i*)^ is given by 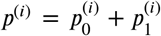, for *i* = 0, 1. The proportion to be estimated is 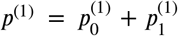, corresponding to the infected individuals, and the^0^ naïve^1^ estimator is biased because the proportion of symptomatic individuals *p*_1_ is overestimated.

### 2.1 Sampling probabilities

For the *j*-th individual in the population (0 < *j* ≤ *N*), define a Bernoulli random variable as follows:

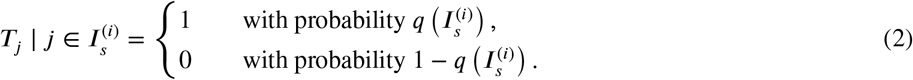

That is, an individual in the category 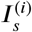 will be tested with probability 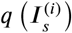, for *s, i =*0, 1.

The sampling probability 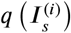 of individuals with symptoms *s* and infection *i* is approximated by

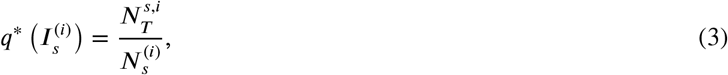

where 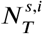 is the number of tested individuals from group 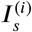. Analogously to (3), *q* (*I*_*s*_), the sampling probability among individuals with symptoms *s*, is approximated by

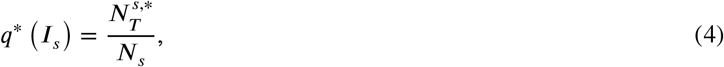

where 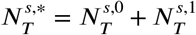. And *q* (*I*^(*i*)^), the sampling probability among individuals with infection status *i*, is approximated by

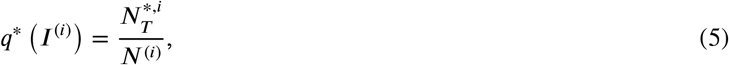

where 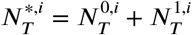. Notice that, except when all symptomatic individuals are sampled (in whose case *N*_1_ is known), the approximations *q*^*^(·) are not estimators of the sampling probabilities because the population values in their denominators are in general unknown. However, when *N* → ∞,

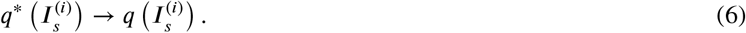

The total of sampled individuals 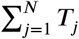 is defined as

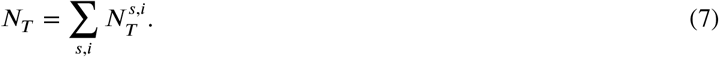

Finally, we define the expected fraction of sampled individuals:

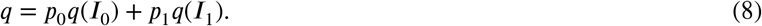

## 3 NO TESTING ERRORS

In case there is no error in testing, the naïve estimator of 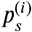 can be naturally defined as

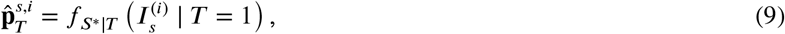

the conditional probability of an individual belonging to the group 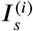, given that they were sampled. The main goal of this paper is to provide a correction for 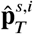, under the assumption that symptomatic individuals are oversampled. A Bayesian approach, inspired from ideas in publication bias, ^6^ leads to

### Proposition 1.

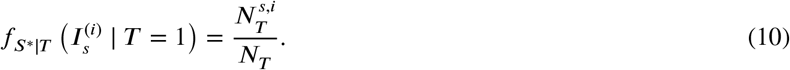

Then 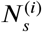, the population size of 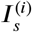, disappears from the sample estimator, and (A1) in the Appendix shows that all information in the sample about the group 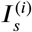 comes from the sampling mechanism 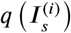. In fact, 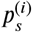 can be seen as the message sent, 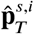 as the message received, and 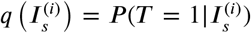 as the channel between them distorting the message. ^11,12^ This interpretation, taken from Shannon’s information diagram, is particularly important to analyze bias as a modification of the information inherent to the prevalence parameter in Section D of the Appendix. ^13^

Analogously to (9) with Proposition 1, the naive estimator of individuals with symptoms *s*, 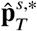, and the naive estimator of individuals with infection status *i*, 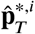, are defined as

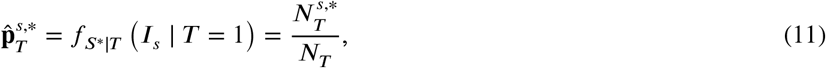

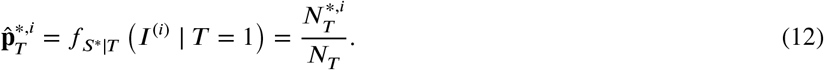

### 3.1 Correction of sampling bias

According to (A1) in the Appendix, some information about the sampling mechanism is needed if any meaningful conclusion is going to be obtained. For the scenario considered in this article, this corresponds to symptomatic individuals being more prone to get tested than asymptomatic ones:

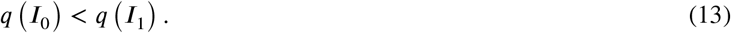

Also corresponding to the intuition that infected and non-infected individuals inside each category are randomly sampled,

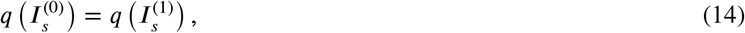

for *s* = 0, 1.

We consider two scenarios. First, when all symptomatic individuals are sampled. Second, building on the previous case, when not all symptomatic are tested, but they are overrepresented in the sample.

#### 3.1.1 All the symptomatic group is sampled

Díaz-Pachón and Rao studied the situation in which, for COVID-19, all symptomatic individuals are tested. ^10^ This scenario corresponds well to some subpopulations like those of universities or industries, in which all symptomatic individuals are required to get tested. In this case, (13) becomes

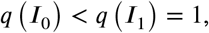

so the proportion *p*_1_ of symptomatic in the population can be fully recovered from the sample as

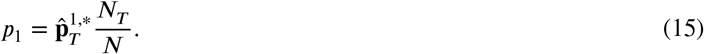

Since, by (14) the sample is assumed to be random among symptomatic individuals,

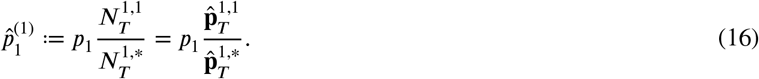

Using now (14) on the asymptomatic group, the prevalence among the asymptomatic is obtained as

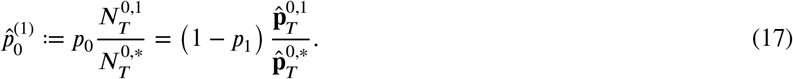

Using (16) and (17), the final sampling-bias corrected prevalence is then taken to be

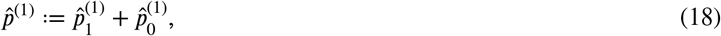

and the random sampling inside each group gives that 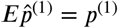, making the estimator unbiased.

Algorithm 1 summarizes the steps from observations to corrected estimate when there is no testing error and all symptomatic individuals are tested.

##### Algorithm 1

Corrected estimator of prevalence without errors and all symptomatic individuals sampled

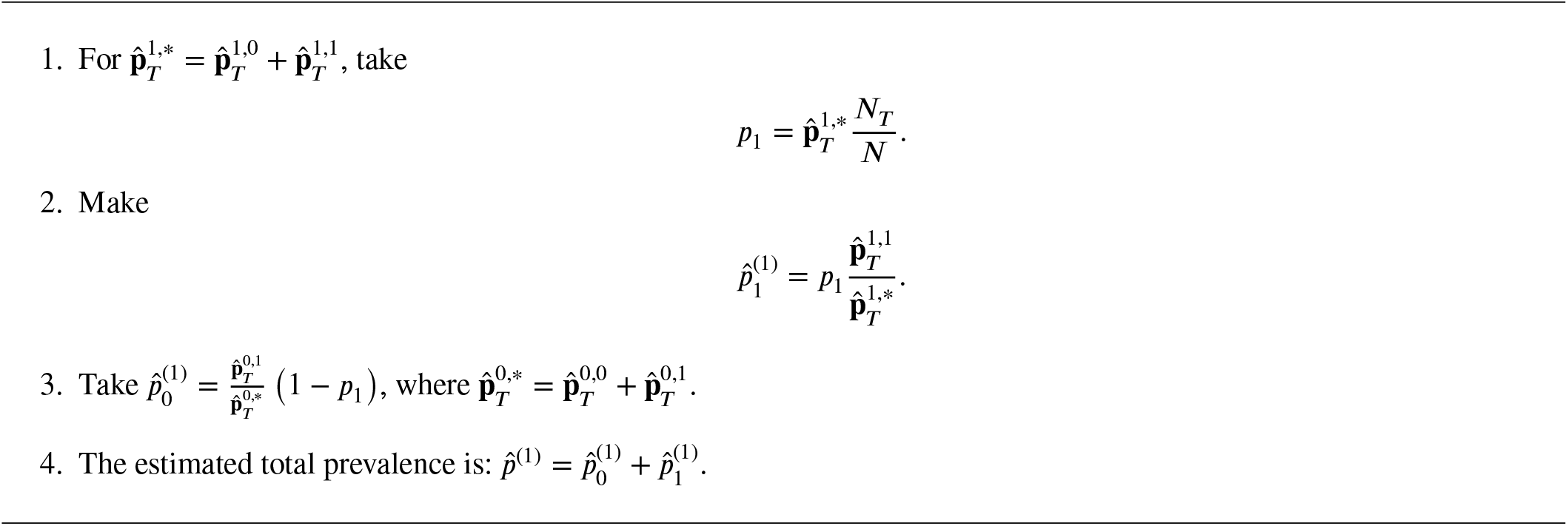

*Remark 1*. Although the scenario of all sampled individuals is also considered by Díaz-Pachón and Rao in the context of COVID-19, ^10^ the correction obtained by Algorithm 1 is stronger than theirs in at least two aspects. First, Díaz-Pachón and Rao considered *m* ≥ 2 categories of symptoms, so that, if *m* is large, an individual with all symptoms is highly likely to be infected; however, with two categories of symptoms, this would correspond to 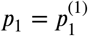, which seems a very strong assumption. Second, Algorithm 1 takes into account the information from the sample to obtain 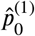 (see step 3 or (17)); instead, Díaz-Pachón and Rao proposed to take *U* uniformly distributed in the interval [0, 1] and make 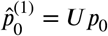.

The following result shows that 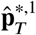 and 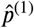 are asymptotically normal. More specifically, it is proven that 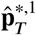 is not a consistent estimator of the true prevalence *p*(1), but 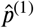 is.

##### Theorem 1.

Suppose *N* → ∞ in such a way that *p*_1_ = *N*_1_*/N* is kept fixed. Then

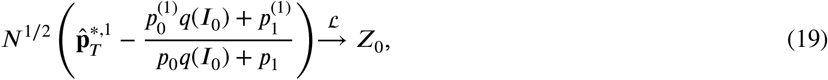

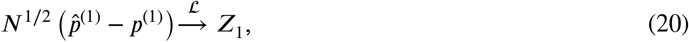

as *N* → ∞, where *Z*_0_ ∼ 𝒩 (0, *V*_01_ + *V*_02_), *Z*_1_ ∼ 𝒩 (0, *V*_03_) are normally distributed, and *V*_01_, *V*_02_, and *V*_03_ are defined in the Appendix.

#### 3.1.2 Not all the symptomatic group is sampled

The main difference between this section and the previous one is that, since now the proportion of symptomatic individuals in the population is not known, it has to be estimated from the sample. Drawing inspiration from cosmological fine-tuning, ^14,15^ the approach will be to use the information in (13) to generate a maximum entropy distribution, which is “the least biased estimate possible on the given information.” ^16^ Next we will use (14) to obtain estimators of prevalence inside each class of symptoms.

##### Theorem 2.

For 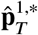, *p*_1_ ∈ (0, 1), *q*^*^ (*I*_0_) < *q*^*^ (*I*_1_) if and only if 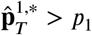.

Theorem 2 shows that, given the basic assumption (13), with high probability *p*_1_ is bounded above by 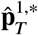. On the other hand, 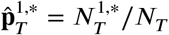 says that there are at least 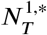 infected symptomatic individuals in the population. Therefore, it makes sense to bound *p*_1_ as follows:

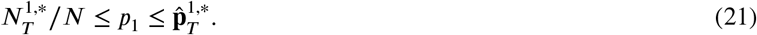

By the maximum entropy principle, ^17^ the corrected estimator of *p*_1_ is taken to be the expectation of a uniform distribution over 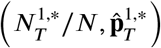. Formally, let *U* be a uniform distribution over the interval 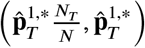. The corrected estimator of *p*_1_ is defined as

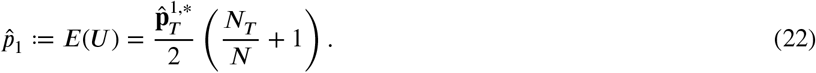

From this point on, we proceed analogously to Subsubsection 3.1.1, replacing *p*_1_ with 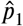 in equations (16), (17), and (18), to obtain

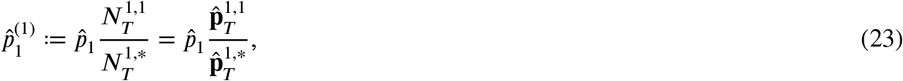

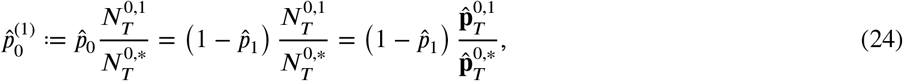

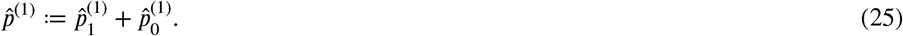

Algorithm 2 summarizes the procedure to produce estimators that correct the sampling bias when there is no testing error and not all symptomatic individuals are sampled.

##### Algorithm 2

Corrected estimator of prevalence without errors and not all symptomatic individuals sampled

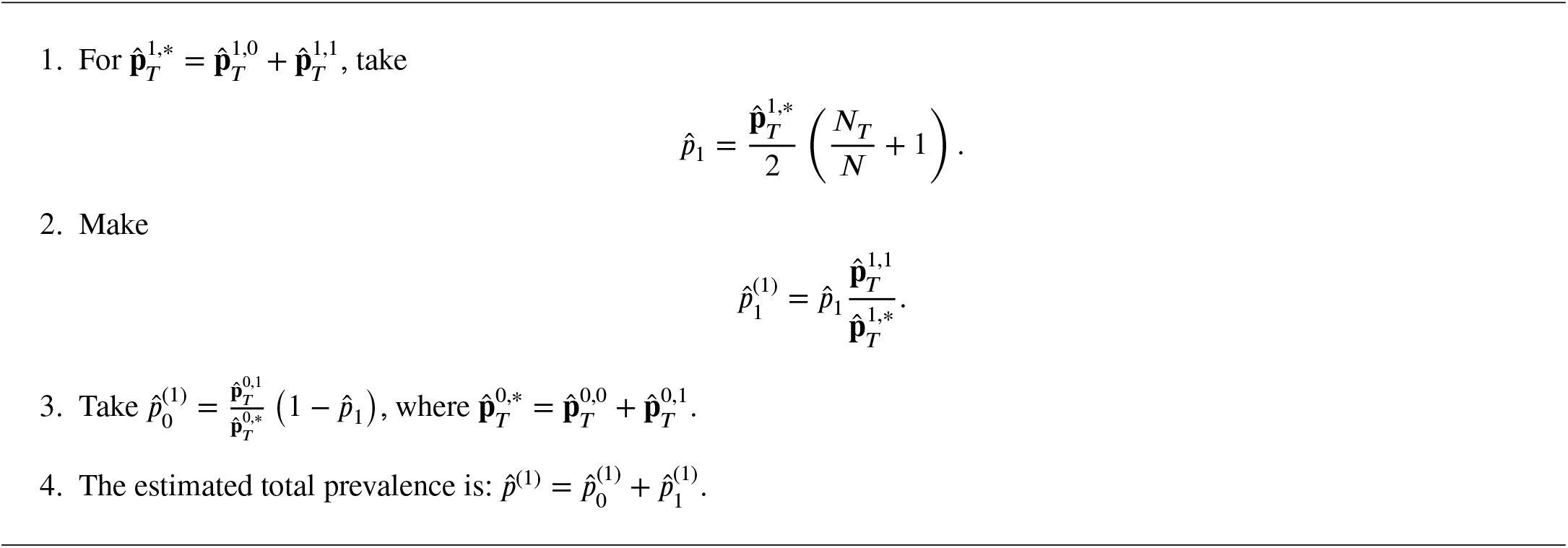

Analogously to the previous asymptotic result, Theorem 3 shows that the naïve and corrected estimators are asymptotically normal. However, once again we find that the naïve estimator is not a consistent estimator of the true prevalence. Moreover, the corrected estimator will only be consistent if 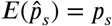, for *s* = 0, 1.

##### Theorem 3.

Suppose *N* → ∞ in such a way that *p*_1_ = *N*_1_*/N* is kept fixed. Suppose additionally that, for *s* = 0, 1, there exists *ρ*_*s*_ ∈ [0, 1], such that

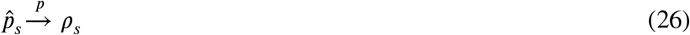

as *N* → ∞, where 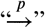 refers to convergence in probability.

Then

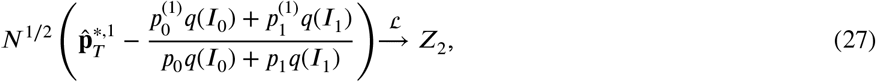

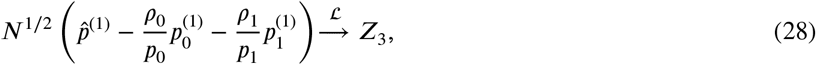

as *N* → ∞, where *Z*_2_ ∼ 𝒩 (0, *V*_11_ + *V*_12_), *Z*_3_ ∼ 𝒩 (0, *V*_13_ + *V*_14_) are normally distributed random variables, and *V*_11_, *V*_12_, *V*_13_, and *V*_14_ are given in the Appendix.

## 4 WITH TESTING ERRORS

When testing errors are considered, the naïve estimators have an additional source of bias. Using (10), in this section, we present first the explicit form of the naïve estimators in the presence of sampling bias and testing errors stratified by symptoms. This is the most general form of naïve estimator considered in this paper. As a corollary, unstratified errors are also considered. With naïve estimators in this general form, we then present their respective corrections.

### Proposition 2.

Let *α*_0_ and *β*_0_ be the false positive and false negative rate for asymptomatic individuals, respectively, and let *α*_1_ and *β*_1_ be the false positive and false negative rate for symptomatic individuals, respectively. The naïve estimators thus become:

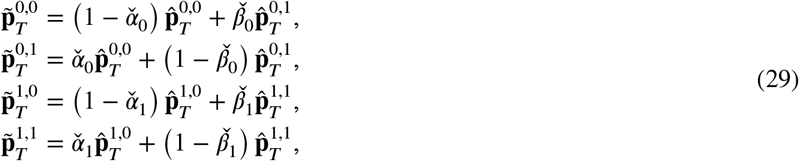

where 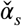 and 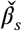, the proportion of false positives and false negatives in the sample, for *s* = 0, 1, approximate *α*_*s*_ and *β*_*s*_ respectively.

Analogously to previous definitions, let 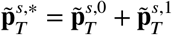 and 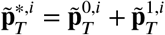.

### Corollary 1.

If errors are not stratified, for *s* = 0, 1, the estimators of Proposition 2,

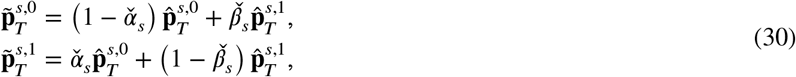

are such that 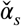 and 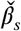, the proportion of false positives and negatives in the sample for symptom class *s*, approximate the probabilities *α* and *β* of false positives and false negatives, respectively, independently of *s*.

*Remark 2*. The right-hand side of (29) and (30) contains the contribution to the naïve estimator by each group in the sample weighted by the probability of their errors. However, in either case, the proportions observed by practitioner are the tilde terms 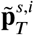 in the left-hand side. The hat terms in the right-hand side, corresponding to (10) are unknown to him.

### 4.1 Correction of testing errors

According to Remark 2, when testing errors are considered, estimators that correct them are necessary before applying the correction to sampling bias. This section presents such estimators.

#### Proposition 3.

For *s* = 0, 1, assume 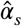 and 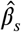 are estimators of *α*_*s*_ and *β*_*s*_, respectively, obtained from different data, satisfying also that they are independent of 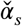 and 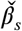. Then the estimators with correction for testing errors

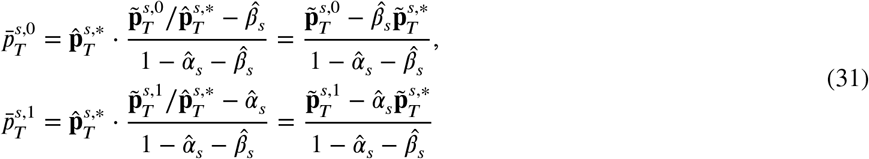

approximate 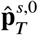 and 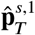 respectively. Moreover, 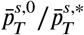 and 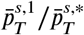 are consistent estimators of 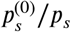 and 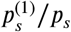, if 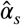 and 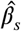 are in turn consistent estimators of *α*_*s*_ and *β*_*s*_, respectively.

#### 4.1.1 All symptomatic group is sampled

When all the symptomatic group is sampled, if 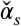 and 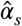 are unbiased estimators of *α*_*s*_, and 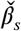 and 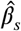 are unbiased estimators of *β*_*s*_, for *s* = 0, 1, following Algorithm 1, we obtain Algorithm 3 substituting 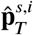 by 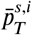.

#### 4.1.2 Not all the symptomatic group is sampled

Analogously to Subsubsection 3.1.2, replace 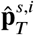 with 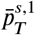 in equations (22)-(25), to obtain

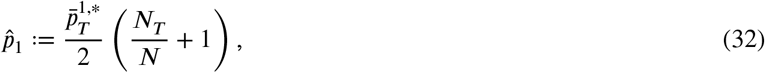

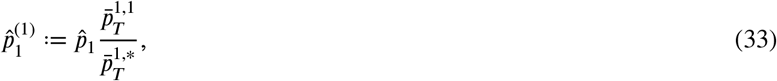

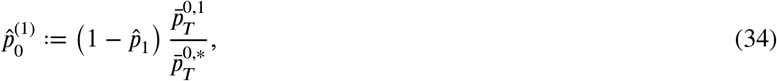

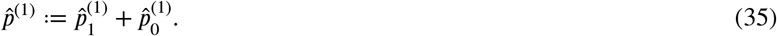

This information can be used to generate Algorithm 4. Theorem 4 summarizes the asymptotic behavior of the estimators involved in this section.

##### Theorem 4.

Suppose the conditions of Theorem 3 hold, and that the estimators prevalences 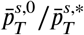 and 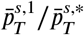 converge to

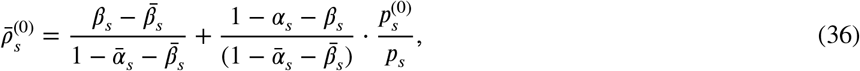

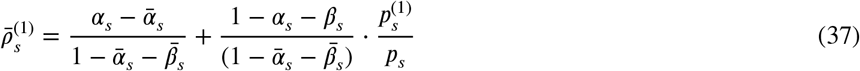

as *N* → ∞. Suppose further that the proportion of individuals wrongly classified in the sample 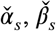, *s* = 0, 1 are independent.

Then

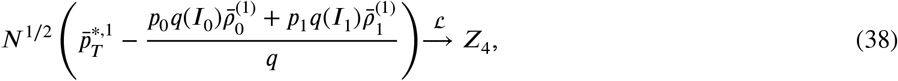

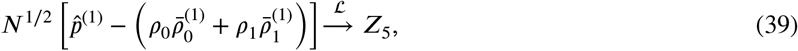

as *N* → ∞, where 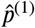 is defined as in (35), *Z*_4_ ∼ 𝒩 (0, *V*_5_ +*V*_6_), *Z*_5_ ∼ 𝒩 (0, *V*_7_ +*V*_8_) are normally distributed random variables, and *V*_5_, *V*_6_, *V*_7_, and *V*_8_ are defined in the Appendix.

*Remark 3*. If stratification is ignored, throughout all this section just take *α* = *α*_0_ = *α*_1_ and 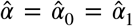. On the other hand, 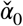 and 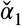 are still distinct quantities, since they correspond to the observed positive error rates of stratum *s* = 0 and *s* = 1 respectively. Then do analogously with the beta terms to obtain *β*, 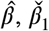 and 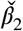.

##### Algorithm 3

Corrected estimator of prevalence with errors and all symptomatic individuals sampled

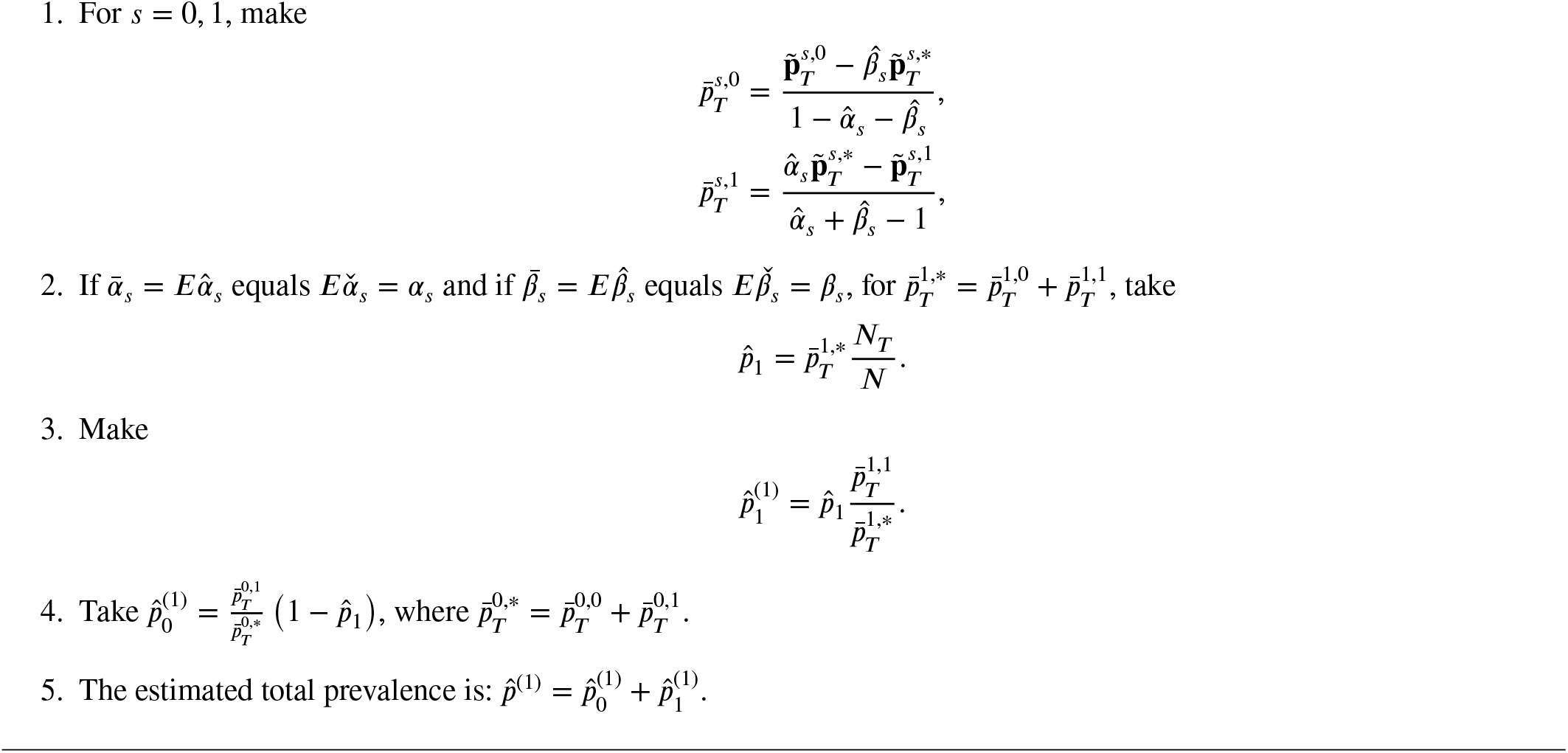

##### Algorithm 4

Corrected estimator of prevalence with errors and not all symptomatic individuals sampled

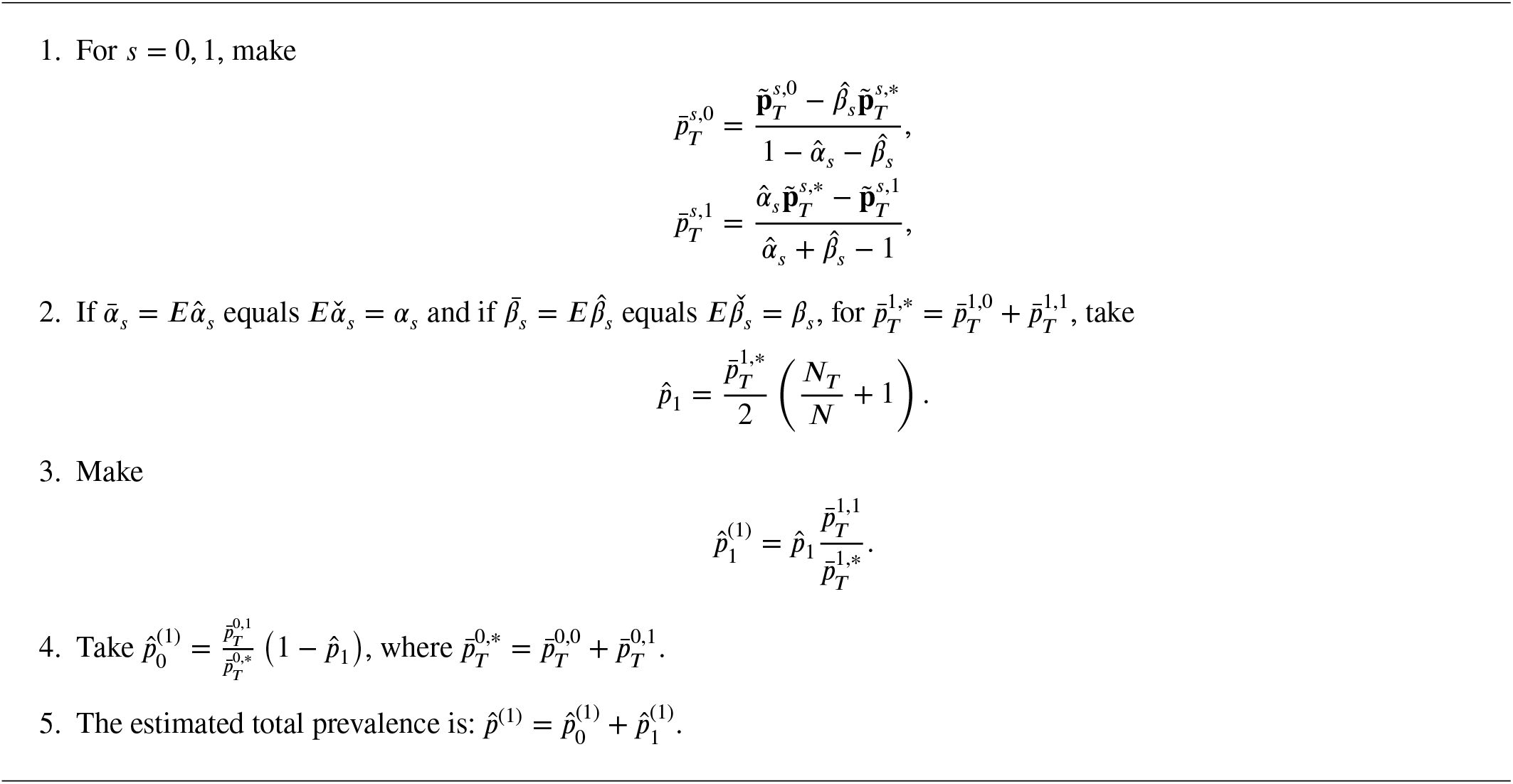

## 5 DATA FROM THE ISRAELI MINISTRY OF HEALTH

In what follows, COVID-19 data from the Israeli Ministry of Health is considered. ^18^ The Ministry of Health publicly released data for individuals tested for COVID-19 via a PCR assay from a nasal swab sample collected between March 22, 2020 and April 7, 2020. The dataset contains information on the test date, test result, clinical symptoms, gender of the individual, known contact with an infected individual and a binary indicator of whether the individual was 60 years of age or older. Symptoms include cough, fever, sore throat, shortness of breath and headache. For the purposes of illustrating the methodology, we will consider this as the population, consisting of 99 232 tested individuals, among whom 1862 were symptomatic (have shortness of breath or have at least three of four symptoms: cough, fever, sore throat, and headache) and 97 370 were asymptomatic. Among the total tested individuals, it was possible to identify 8393 infections through PCR testing. Among the individuals who tested positive, 1754 were symptomatic. The characteristics of the data set are presented in Table 2.

**TABLE 2.** Observed disease status by category of symptoms.

|  | Positive | Negative | Total |
| --- | --- | --- | --- |
| Symptomatic | 1 754 | 108 | 1 862 |
| Asymptomatic | 6 639 | 90 731 | 97 370 |
| Total | 8 393 | 90 839 | 99 232 |

Error rates will be stratified by symptoms. Thus, let *α*_0_ and *α*_1_ be the false positive rate for asymptomatic and symptomatic individuals, respectively, and *β*_0_ and *β*_1_, the false negative rate for asymptomatic and symptomatic individuals, respectively. Although we do not have exact numbers of stratified false-positive and false-negative rates for PCR tests, a public report from UK Government Office for Science in 2020 indicated that the median false positive rate in the UK’s COVID-19 RT-PCR testing program is 2.3% with IQR of 0.8% to 4.0%. ^19^ Moreover, Arévalo-Rodríguez et al stated that after they collected information among all patients from 34 studies, the summary estimate of the false-negative rate was 13% with range of 1.8% to 58%. ^20^ For the purpose of investigating how the estimates change with different assumed error rates, we will compare different combinations of error rates within the reasonable ranges according to the literature we found. The combinations of error rates in the population are assumed to have the values in (40). The actual number of individuals inside each group can be found in Table 3 after correcting Table 2 for these errors.

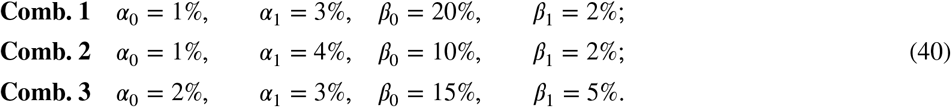

The real prevalence with **Comb. 1** is then

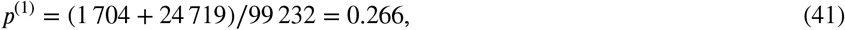

and prevalence among the asymptomatic is 24 719*/*97 370 = 0.254.

The real prevalence with **Comb. 2** is then

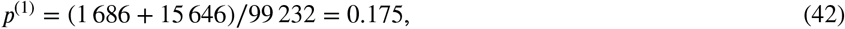

and prevalence among the asymptomatic is 15 646*/*97 370 = 0.161.

The real prevalence with **Comb. 3** is then

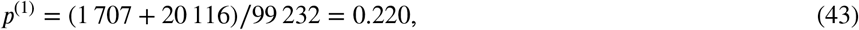

and prevalence among the asymptomatic is 20 116*/*97 370 = 0.207.

Additionally, in (44) we assume some sample error rates for the combinations (40). We emphasize that the actual values of (44) are “known unknowns” to the practitioner, ^21^ and it is precisely their effect what needs to be corrected.

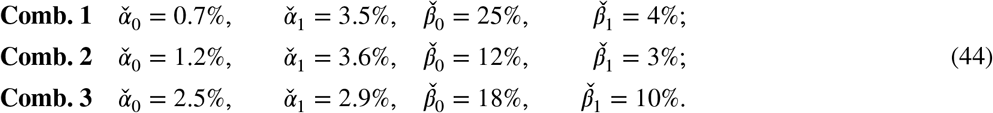

Finally, active information (defined in Appendix C) is used to compare how well Algorithms 3 and 4 and other proposed estimators in the literature are doing with respect to the real prevalence. The best estimator will be the one with active information *Î*^+^ closer to 0. The competitors will be the method proposed by Díaz-Pachón and Rao, which assumes all symptomatic individuals are sampled, correcting only for sample bias and ignoring testing errors; ^10^ Diggle’s Bayesian approach, which corrects for imperfect testing but ignores sampling bias; ^22^ and the Rogan-Gladen estimate, a frequentist method that only corrects for testing errors too. ^23^ Neither of the competitors corrects for sampling bias and testing errors at the same time. In fact, as much as we searched, we could not find a methodology that simultaneously corrects for imperfect testing and sampling bias; this will be reflected in the analysis. All of the results are presented in Table 8.

Each of the following protocols presents a table with the sample results. These correspond to the observations given by (29). These values, the population size, and the estimated error rates from a different study (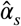 and 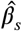) will be the input of an R program, for which the code is available at https://github.com/kalilizhou/BiasCorrection.git, with the four algorithms proposed in this article. The program thus obtains the correction. As a simplifying assumption in the remaining of this section, we take 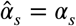 and 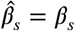, for *s* = 0, 1.

**TABLE 3.**
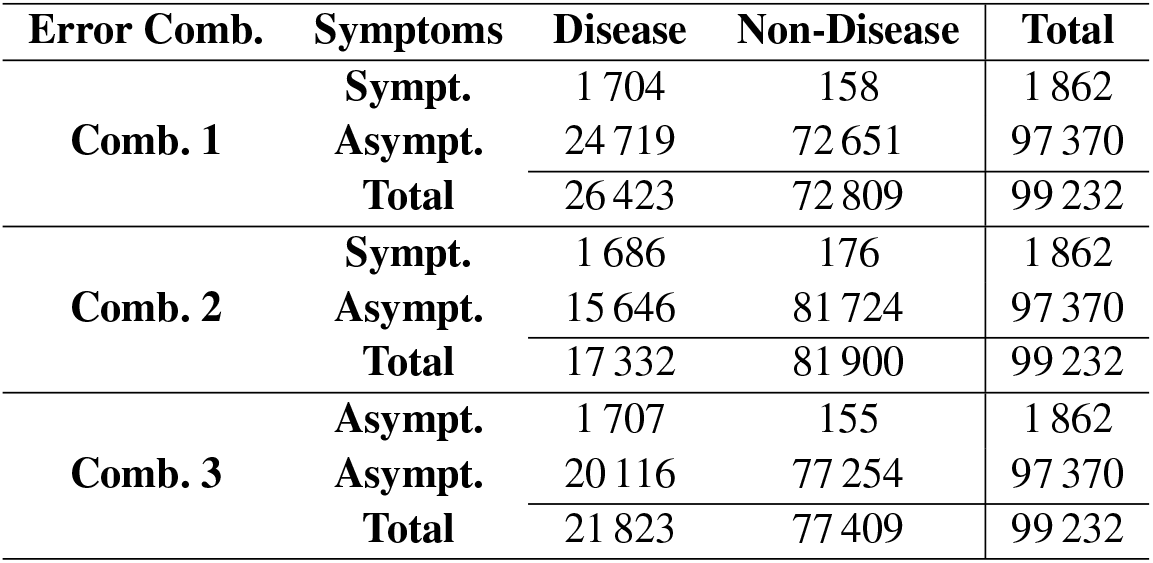
Real proportions under stratified errors with different error combinations.

| Error Comb. | Symptoms | Disease | Non-Disease | Total |
| --- | --- | --- | --- | --- |
| **Comb. 1** | **Sympt.** | 1 704 | 158 | 1 862 |
|  | **Asympt.** | 24 719 | 72 651 | 97 370 |
|  | **Total** | 26 423 | 72 809 | 99 232 |
| **Comb. 2** | **Sympt.** | 1 686 | 176 | 1 862 |
|  | **Asympt.** | 15 646 | 81 724 | 97 370 |
|  | **Total** | 17 332 | 81 900 | 99 232 |
| **Comb. 3** | **Sympt.** | 1 707 | 155 | 1 862 |
|  | **Asympt.** | 20 116 | 77 254 | 97 370 |
|  | **Total** | 21 823 | 77 409 | 99 232 |

### Sampling Protocol 1

In the first scenario, all symptomatic individuals are sampled, as considered by Díaz-Pachón and Rao. ^10^ The sample consists of 2483 individuals. Among these, 1862 (75%) are symptomatic and 621 (25%) are asymptomatic. The sample error rates are taken from (44). The observed sampling results, corresponding to (29), are given by Table 4.

**TABLE 4.** Stratified observed sample for 75% symptomatic and 25% asymptomatic, with all symptomatic individuals sampled.

| Error Comb. | Symptoms | Positive | Negative | Total |
| --- | --- | --- | --- | --- |
| **Comb. 1** | Symptomatics | 1 641 | 221 | 1 862 |
|  | Asymptomatics | 122 | 499 | 621 |
|  | Total | 1 763 | 720 | 2 483 |
| **Comb. 2** | Symptomatics | 1 642 | 220 | 1 862 |
|  | Asymptomatics | 94 | 527 | 621 |
|  | Total | 1 736 | 747 | 2 483 |
| **Comb. 3** | Symptomatics | 1 541 | 321 | 1 862 |
|  | Asymptomatics | 117 | 503 | 620 |
|  | Total | 1 658 | 824 | 2 482 |

According to Table 4, for instance with **Comb. 1**, the naïve estimator is 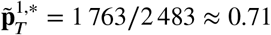. Since all the symptomatic group is sample, we use Algorithm 3, which produces the corrected estimator 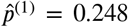. Table 8 presents these results as well as those of the other methods.

In this case, Diggle’s correction was not implemented because it involves combinations in its logarithm that are difficult to approximate when the sample is moderately large. Under the assumption of sampling all symptomatic individuals, Díaz-Rao works very well, and RGE performs as poorly as the naïve estimators. Our Algorithm 3 is the best correction to the the naïve estimate, producing the closest-to-zero actinfo. The corrected estimators of prevalence obtained from Algorithm 3 almost equal the real prevalence for all combinations of testing errors.

For the next protocols, the assumption that all symptomatic individuals were sampled is removed, which implies that the Diaz-Rao correction cannot be assessed and Algorithm 4 is followed.

### Sampling Protocol 2

The sample consists of 200 individuals. Among these, 150 (75%) are symptomatic and 50 (25%) are asymptomatic. For both the symptomatic and asymptomatic groups, the sampling proportions are taken according to Table 3. Table 5 shows the observed sample. The summary of results under different methods is shown in Table 8.

**TABLE 5.** Stratified observed sample for 75% symptomatic and 25% asymptomatic (not all symptomatic individuals sampled).

| Error Comb. | Symptoms | Positive | Negative | Total |
| --- | --- | --- | --- | --- |
| Comb. 1 | Sympt. | 132 | 18 | 150 |
|  | Asympt. | 10 | 40 | 50 |
|  | Total | 142 | 58 | 200 |
| Comb. 2 | Sympt. | 132 | 18 | 150 |
|  | Asympt. | 8 | 42 | 50 |
|  | Total | 140 | 60 | 200 |
| Comb. 3 | Sympt. | 125 | 25 | 150 |
|  | Asympt. | 9 | 41 | 50 |
|  | Total | 134 | 66 | 200 |

Table 8 shows that, in this sampling scenario, our Algorithm 4 still has the best performance. In fact, Diggle’s and Rogan-Gladen’s estimates do as poorly as the naïve estimate. Algorithm 4 beats its competitors because it is the only one that corrects for sampling bias, whereas the other two only correct for testing errors. Notice that the additional information of Protocol 1 (knowing that all symptomatic individuals were sampled), in comparison to Protocol 2, greatly improves the performance of the correction, as reflected by the active information.

### Sampling Protocol 3

In this scenario there are 100 symptomatic and 100 asymptomatic individuals. Again, the proportions inside each group were taken from Table 3. The observed sample is given in Table 6. After correcting the estimates, the summary of results under different methods for this sampling protocol is also presented in Table 8.

**TABLE 6.** Stratified observed sample for 50% symptomatic and 50% asymptomatic.

| Error Comb. | Symptoms | Positive | Negative | Total |
| --- | --- | --- | --- | --- |
| Comb. 1 | Sympt. | 89 | 11 | 100 |
|  | Asympt. | 19 | 81 | 100 |
|  | Total | 108 | 92 | 200 |
| Comb. 2 | Sympt. | 89 | 11 | 100 |
|  | Asympt. | 15 | 85 | 100 |
|  | Total | 104 | 96 | 200 |
| Comb. 3 | Sympt. | 83 | 17 | 100 |
|  | Asympt. | 19 | 81 | 100 |
|  | Total | 102 | 98 | 200 |

Compared to Protocol 2, Protocol 3 has less sampling bias. Therefore, all the methods perform better than in the previous scenario. But Algorithm 4 still works better than competitors.

### Sampling Protocol 4

This sample is truly random, with *N*_*T*_ = 200, and it is obtained from Table 3. The observed sample is presented in Table 7. The results of the different methods for this scenario are presented in Table 8.

**TABLE 7.** Stratified observed random sample of size 200.

| Error Comb. | Symptoms | Positive | Negative | Total |
| --- | --- | --- | --- | --- |
| Comb. 1 | Sympt. | 3 | 1 | 4 |
|  | Asympt. | 39 | 157 | 196 |
|  | Total | 42 | 158 | 200 |
| Comb. 2 | Sympt. | 3 | 1 | 4 |
|  | Asympt. | 30 | 166 | 196 |
|  | Total | 33 | 167 | 200 |
| Comb. 3 | Sympt. | 3 | 1 | 4 |
|  | Asympt. | 37 | 159 | 196 |
|  | Total | 40 | 160 | 200 |

**TABLE 8.** Comparison among methods of all sampling protocols with different combinations of stratified error rates.

| Corrected Estimates $\hat{p}^{(1)}$ (Active Information $\hat{I}^+$) | | | | | | | |
| --- | --- | --- | --- | --- | --- | --- | --- |
| Error Comb. | Real Prevalence | Sampling Protocol | Naïve | Díaz-Rao | Diggle | Rogan-Gladen | Algorithms 3 and 4 |
| Comb. 1 | **p = 0.266** | 1 | 0.710 (0.981) | 0.212 (-0.229) | - | 0.729 (1.009) | **0.248 (-0.069)** |
|  |  | 2 | 0.710 (0.981) | - | 0.735 (1.016) | 0.729 (1.009) | **0.486 (0.602)** |
|  |  | 3 | 0.540 (0.707) | - | 0.535 (0.699) | 0.564 (0.751) | **0.398 (0.401)** |
|  |  | 4 | 0.210 (-0.237) | - | 0.003 (-4.667) | **0.249 (-0.066)** | 0.244 (-0.086) |
| Comb. 2 | **p = 0.175** | 1 | 0.699 (1.387) | 0.167 (-0.045) | - | 0.711 (1.402) | **0.173 (-0.011)** |
|  |  | 2 | 0.700 (1.388) | - | 0.695 (1.379) | 0.712 (1.403) | **0.441 (0.926)** |
|  |  | 3 | 0.520 (1.091) | - | 0.493 (1.035) | 0.530 (1.109) | **0.344 (0.679)** |
|  |  | 4 | 0.165 (-0.057) | - | 0.003 (-4.248) | 0.173 (-0.014) | 0.168 (-0.047) |
| Comb. 3 | **p = 0.220** | 1 | 0.668 (1.111) | 0.204 (-0.076) | - | 0.701 (1.159) | **0.215 (-0.019)** |
|  |  | 2 | 0.670 (1.114) | - | 0.690 (1.143) | 0.703 (1.161) | **0.448 (0.713)** |
|  |  | 3 | 0.510 (0.841) | - | 0.503 (0.826) | 0.537 (0.892) | **0.371 (0.524)** |
|  |  | 4 | 0.200 (-0.095) | - | 0.003 (-4.477) | **0.215 (-0.024)** | 0.209 (-0.050) |

In this scenario, without sampling bias, all estimates perform extremely well except Diggle’s estimate. Diggle’s Bayesian estimates grossly overcorrect to the point of removing more than 4 nats of information with respect to the real prevalences. On the other hand, Rogan-Gladen’s frequentist estimates are optimal, and Algorithm 4 work quite well. The results of these two methods are very close to the real prevalence.

Table 8 summarizes the results of all protocols and all combinations of testing errors. Our proposed Algorithms 3 (first row in each combination) and 4 (rows 2-4 in each combination) are always the best in Sampling Protocols 1-3 and perform as well as the naïve estimator under random sampling (Protocol 4), according to the actinfo assessment. The result shows the promising ability of our proposed algorithms to correct for both sampling bias and testing errors in prevalence estimation.

## 6 DISCUSSION

Timely and accurate prevalence estimation of a disease is one of the most fundamental concepts in epidemiology and its importance is because it provides a measure of disease burden in a population at a particular point in time. It can also be part of a compendium of measures used to inform public health prevention policies to help slow the spread of disease through the population. To provide prevalence estimates that are reliable and generalizable, the sample must be comprehensive enough to capture all relevant subpopulations in the general population and as mentioned, for a number of diseases this can be challenging because many of these sub-populations can be hard-to-reach. Thus, sampling bias corrections are needed. Interestingly, this paper has presented new methodology where biased samples result due to over-sampling of symptomatic individuals. Such biased samples are here shown to be inconsistent in terms of not converging to the true proportion of infected individuals. In addition, Algorithms 1, 2, 3, and 4 go further and present corrections both for sampling bias and testing errors. Such corrections either eliminate bias completely (Algorithms 1 and 3), or reduce it substantially (Algorithms 2 and 4) when testing error rates are known or can be estimated consistently. However, the methodology generalizes easily regardless of how the biased samples resulted.

A limitation of our study is that we do not estimate error rates directly from our sample, but take the estimator from a previous independent sample. If this is not the case, then at least under the random sampling situation, prevalence can still be estimated using a Bayesian approach described by Diggle. ^22^ This naturally results in increased variability of the prevalence estimate and relies on a reasonable prior distribution being elicited for the prevalence.

Sample pooling has also been proposed as an efficient way to estimate population prevalence because if the disease prevalence is low, then little information is accrued from individual tests. ^24^ This is sometimes called group testing. However, this implicitly assumes random sampling of pools which is clearly not the case considered here.

Another approach is to use population seroprevalence complex surveys. ^25,26^ While inherently much more difficult to conduct and analyze, these can also suffer from non-ignorable non-response which can lead to biased estimates of prevalence. Indeed, biased sampling can be more generally cast within a missing data framework and the impact of different missing data mechanisms has been studied. ^27^

For some diseases it is becoming more common to use administrative data to estimate disease prevalence since for many countries these data cover large proportions of the population. Examples include Canada, Denmark and Italy among others. This requires some effort to properly assemble these data sources, ^28^ but they have to date not proven as useful for emerging diseases like COVID-19 where surveillance studies dominated the earlier days of the pandemic.

In First-World countries, particularly in urban areas, testing practices seem to be well-described by oversampling of symptomatic individuals, sometimes even testing the whole group in a subpopulation, as it is the case with COVID-19 testing in universities and industries. A possible extension, however, is to consider the opposite situation in which the symptomatic group is under-sampled, producing an estimator that is biased because it underestimates prevalence. Such scenario is certainly relevant for COVID-19 too in several Third World countries, and even in difficult-to-reach subpopulations of First World Countries.

## Data Availability

Code to implement Algorithm 1 is available at https://github.com/kalilizhou/BiasCorrection.git. The data used in Section 6 is publicly available at https://github.com/nshomron/covidpred.

https://github.com/nshomron/covidpred

## Author contributions

D. A. D. P. and J. S. R. conceptualized the methodology framework and the paper. L. Z., D. A. D. P., and O. H. developed the methodology details. L. Z. ran the examples and produced the R code. J. S. R. and C. Z. ran the simulations. D. A. D. P., J. S. R., and O. H. reviewed and proofread the paper.

## Financial disclosure

D.A.D.P., C.Z., and J.S.R. acknowledge the support of the Copeland Foundation Award 2022 from the Department of Public Health Sciences at the University of Miami.

## Conflict of interest

The authors declare no potential conflict of interests.

## Data availability statement

Code to implement Algorithms 1-4 is available at https://github.com/kalilizhou/BiasCorrection.git. The data used in Section 5 is publicly available at https://github.com/nshomron/COVIDpred.

## Acknowledgements

The authors thank the comments from the two anonymous reviewers that greatly improved the quality and readability of this manuscript.

**How to cite this article:** Zhou L., Díaz-Pachón D. A., Zhao C., and Rao J. S. (2022), Correcting prevalence estimation for biased sampling with testing errors,, *2022;00:1–13*.

## APPENDIX

### A GENERAL PROOFS

*Proof of Proposition 1*.

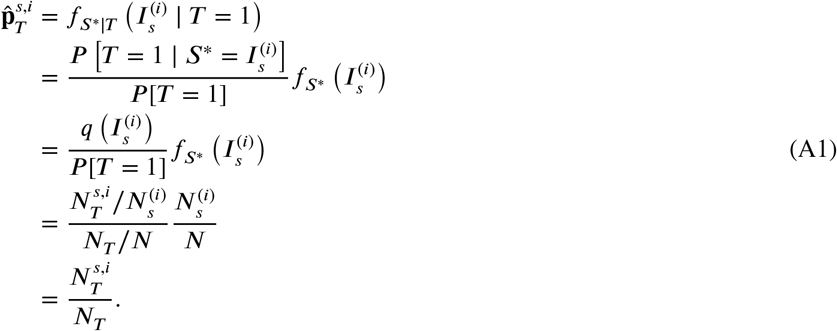

Remember that terms without subindex *T* are here population values, whereas terms with the subindex *T* are sample values. Notice in the fourth and fifth steps that 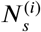 cancels. Therefore, all the remaining information about 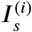 comes from 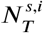, the number of tested (sampled) individuals with symptoms *s* and infectious status *i*. Now, from the third equality, this value is seen to come from 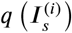, the sampling probability of the group 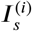. Therefore, all knowledge of 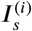 comes from whatever knowledge we have about the sample mechanism 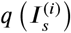.

□

*Proof of Theorem 2*.

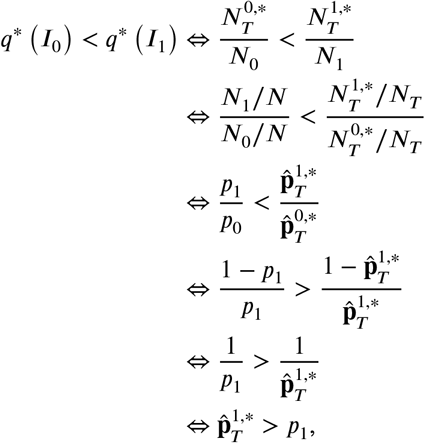

where the fourth step used that 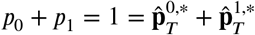.

□

*Proof of Proposition 2*. This proposition follows directly from the definition of testing errors. Consider for instance the first equation of (29). It stipulates that whereas the proportion of sampled individuals with *s* = *i* = 0 is 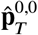, the reported fraction 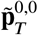 of sampled individuals with *s* = *i* = 0 differs from 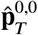 by an amount 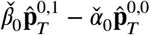, where the first term 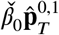 is the fraction of (0, 1)-individuals wrongly classified as (0, 0), whereas the second term 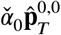 is the fraction of (0, 0)-individuals wrongly classified as (0, 1). The other three equations of (29) are motivated analogously.

□

*Proof of Proposition 3*. It follows from equations (29) and (31) that 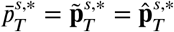 for *s* = 0, 1. This, and another application of (29) and (31) gives

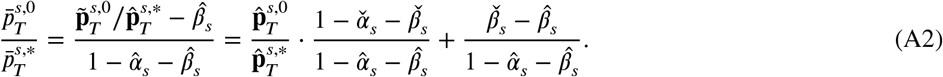

Since 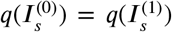 by (14), it follows that 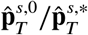 is a consistent estimator of 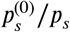, and by assumption, 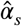 and 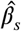 are consistent estimators of *α*_*s*_ and *β*_*s*_ respectively. Moreover, Lemma 2 below implies that 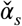 and 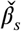 are consistent estimators of *α* and *β* as well. From this and (A2) it follows that 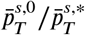 is a consistent estimator of 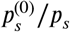. The fact that 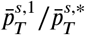 is a consistent estimator of 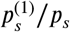 is proved in the same way.

□

### B ASYMPTOTICS

As a preparation, we prove the following lemma that will be used as assumption in the main result of this section:

#### Lemma 1.

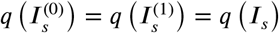.

*Proof*. The first equality is obtained by assumption (14). In order to prove the second equality, we use that a randomly chosen individual from *I*_*s*_ belongs to 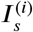 with probability 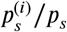 for *i* = 0, 1. Conditioning on which subcohort of *I*_*s*_ the individual belongs to, it follows that

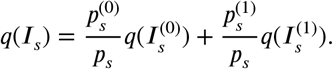

Therefore, since *q*(*I*_*s*_) is a weighed average of 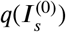 and 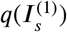, the second equality of Lemma 1 follows from the first one.

□

Hössjer et al. (2023) proved a couple of theorems that we will use to prove the asymptotic results for the estimators discussed in this paper. ^27^ We present them here for completeness, fitting their notation to ours.

#### Theorem 5

(Theorem 1 of Hössjer et al. (2023)). Let *N* → ∞ in such a way that *N*_1_/*N* is always fixed, that Lemma 1 holds for fixed *q*(*I*_0_) and *q*(*I*_1_), and that there exists *ρ*_*s*_ ∈ [0, 1] such that 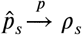, as *N* → ∞, for *s* = 0, 1. Then

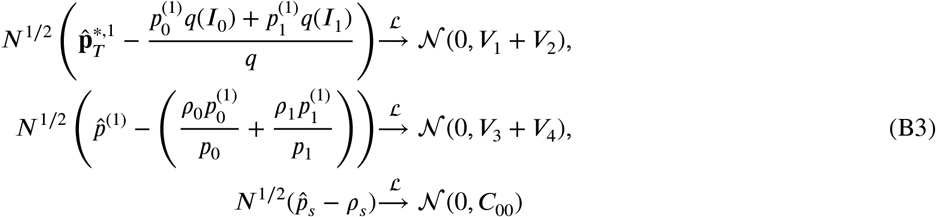

as *N* → ∞, where *q* is defined in (8),

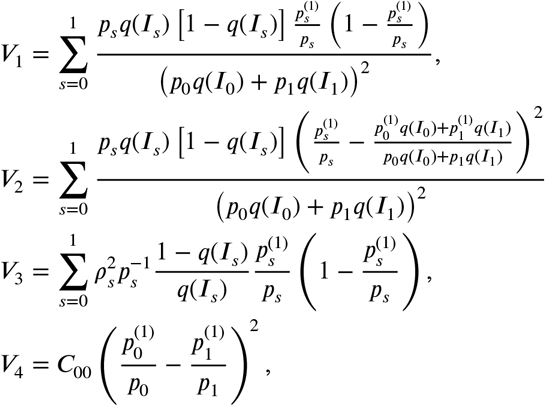

and

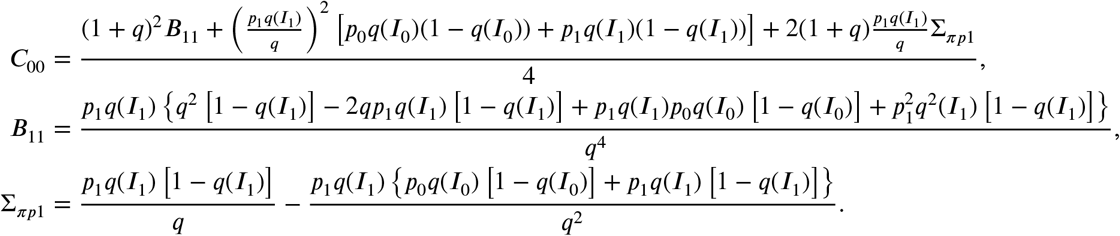

*Proof of Theorem 1*. Since 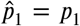 and 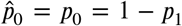 are known, convergence in probability 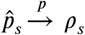 trivially holds with *ρ*_*s*_ = *p*_*s*_, whereas the asymptotic weak limit of 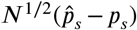 in (B3) degenerates to a one point distribution at 0 (*C*_00_ = 0). Since *q*(*I*_1_) = 1, *V*_1_, *V*_2_, and *V*_3_, in Theorem 5 are readily simplified to

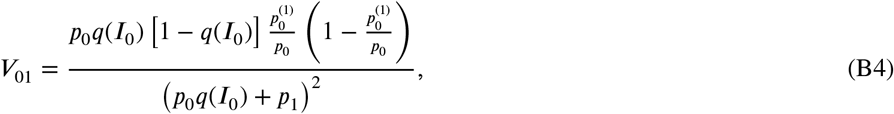

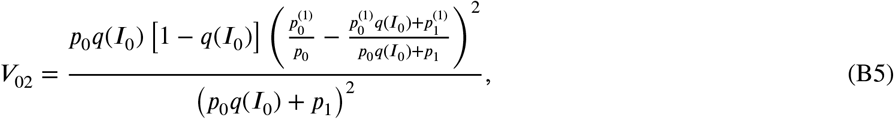

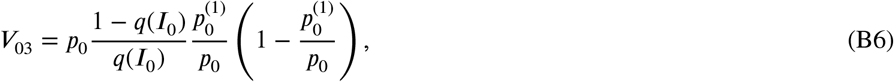

whereas *V*_4_ = 0 since *C*_00_ = 0.

□

*Proof of Theorem 3*. It is obtained directly from Theorem 5 by substituting *V*_1_, *V*_2_, *V*_3_, and *V*_4_ for *V*_11_, *V*_12_, *V*_13_, and *V*_14_, respectively, where

@
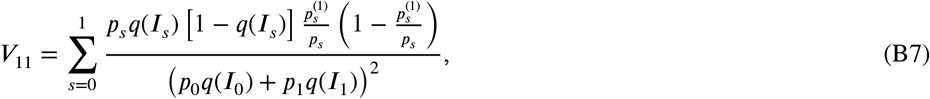

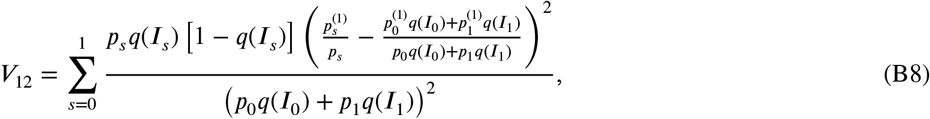

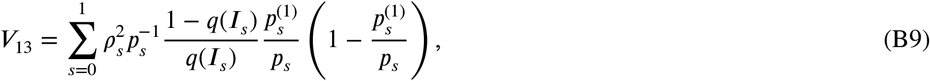

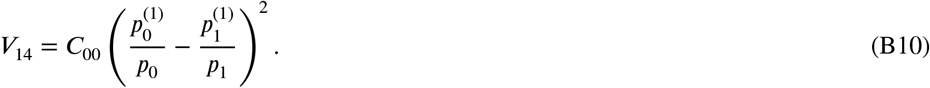

□

#### B.1 Asymptotics with testing errors

Before proving the asymptotic results with testing errors, some previous assumptions and results are used. First, we assume that the existing estimators of error rates are asymptotically normal. That is, there exists 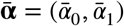 and 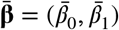 such that, for 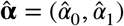 and 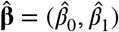,

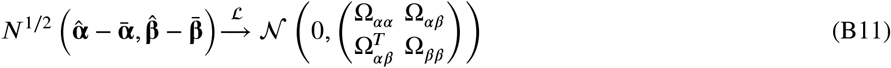

as *N* → ∞, where each of the terms in the variance-covariance matrix is a 2 × 2 matrix, and

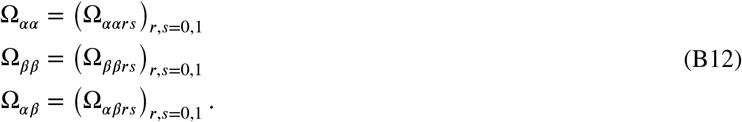

We also assume that 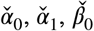, and 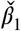 are all independent. Moreover, the following Lemma will be used:

##### Lemma 2

(Lemma 2 from Hössjer et al. (2023)). The proportions of false positive and negatives in the sample satisfy

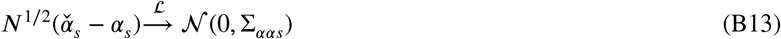

and

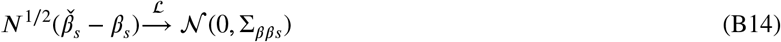

respectively as *N* → ∞, with

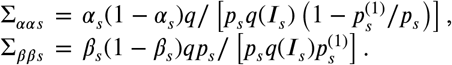

After transforming the notation of Hössjer et al. (2023) to our notation, the asymptotic results of Theorem 4 are a direct consequence of the following result from Hössjer et al. (2023):

##### Theorem 6

(Theorem 2 of Hössjer et al. (2023)). Suppose the conditions of Theorem 3 hold, and additionally that the estimators 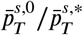 and 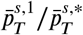 of prevalences of unaffected and affected in symptom group *s* converge to

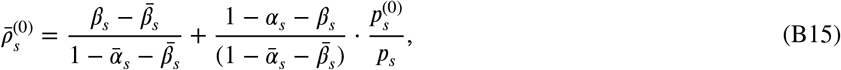

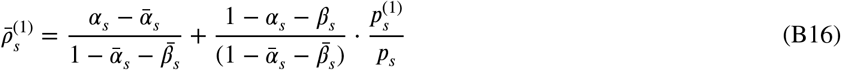

respectively as *N* → ∞. Suppose further that the proportion of individuals wrongly classified in the sample 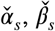 *s* = 0, 1 are independent. Then

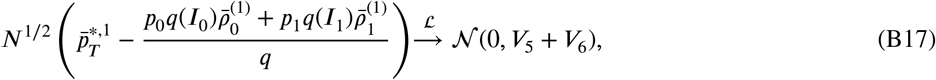

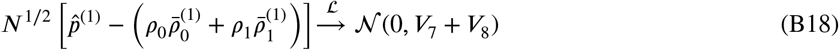

as *N* → ∞, where 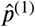 is defined as in (35),

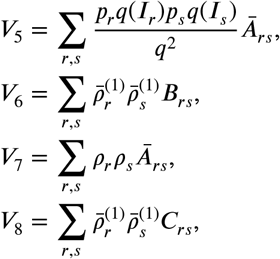

whereas

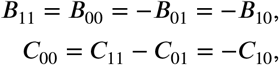

are defined in Theorem 5. Moreover,

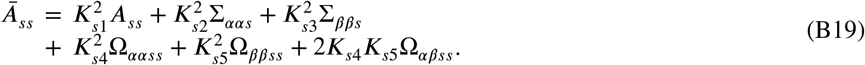

and

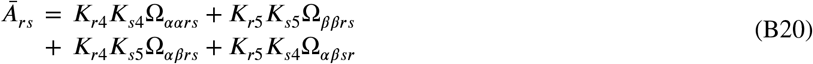

when *r* ≠ *s*, with

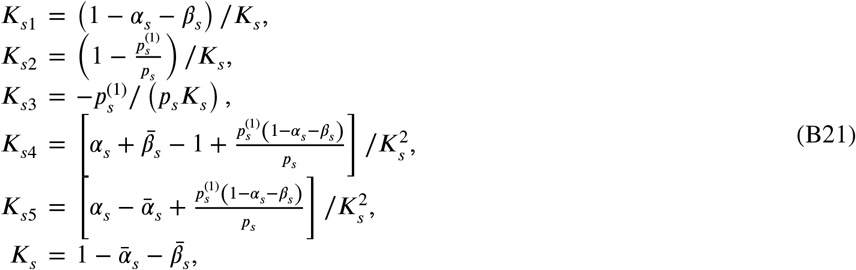

and finally

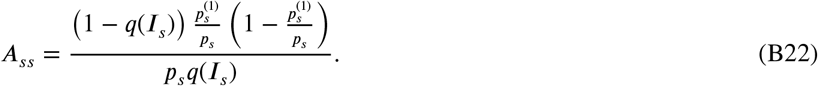

Notice that, if 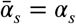 and 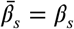 (that is, if the error rates are estimated consistently), then 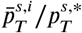 is a consistent estimator of 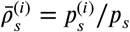 for *i* = 0, 1. In particular, 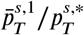 is a consistent estimator of the prevalence 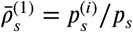 among the individuals of symptom group *s*.

### C ACTIVE INFORMATION: THE INDEX

Active information (actinfo) was introduced in search problems to quantify the amount of Shannon information introduced by the programmer in a search problem. ^29,30,31^ In machine learning, it has been used to show that no algorithm performs well for a large class of problems, in agreement with the so-called No Free Lunch Theorems. ^32,33,34^ It has also been used for mode hunting, ^35,36^, and to compare neutral to non-neutral models in population genetics. ^37^

We now use active information to analyze the bias. Through the eyes of actinfo, the bias is formally seen as the addition (if the parameter is overestimated) or subtraction (if the parameter is underestimated) of relevant information in the estimation of the parameter. Formally, active information is defined as

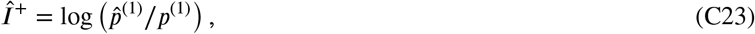

where the logarithm is taken to be in base *e*, so that information is measured in nats. Thus defined, active information measures the amount of Shannon information of the estimator 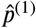 to the true proportion *p*_(1)_, and it is the quantity that is averaged in the Kullback-Leibler divergence. _38_ That is, if the true proportion is overestimated, the active information will be positive and large; if the true proportion is underestimated, the active information will be negative; and if the true proportion is accurately estimated, the active information will be around zero. _39,40_ Because of Theorem 4, we interpret (C23) as an approximation of 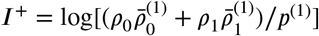.

### D SIMULATION

This section uses simulation to analyze the asymptotic behavior of the corrected estimator. The population has the following features:

- The proportion of positive cases with symptoms 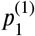 is 15%,
- the proportion of negative cases with symptoms 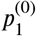 is 5%,
- the proportion of positive cases without symptoms 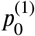 is 5%,
- and proportion of negative cases without symptoms 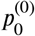 is 75%.

Thus, the prevalence is 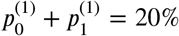, the proportion of symptomatic individuals in the population is 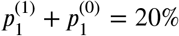, so the proportion of asymptomatic in the population is 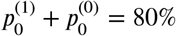.

#### D.1 Correction without testing errors

We will run the simulation for multiple proportions of asymptomatic individuals getting sampled and will compare the results using a boxplot. The true prevalence will be known in the simulation, allowing us to evaluate the accuracy of our estimators.

##### D.1.1 All the symptomatic group is sampled

Assuming a population size of 10 000, we initially sampled all symptomatic individuals. However, we increased the sample size by including more asymptomatic individuals as we changed the proportion of those getting tested. This resulted in both the corrected and naive estimators approaching the true value, and the variance of the estimators decreasing, as shown in Figure D1.

**FIGURE D1.**
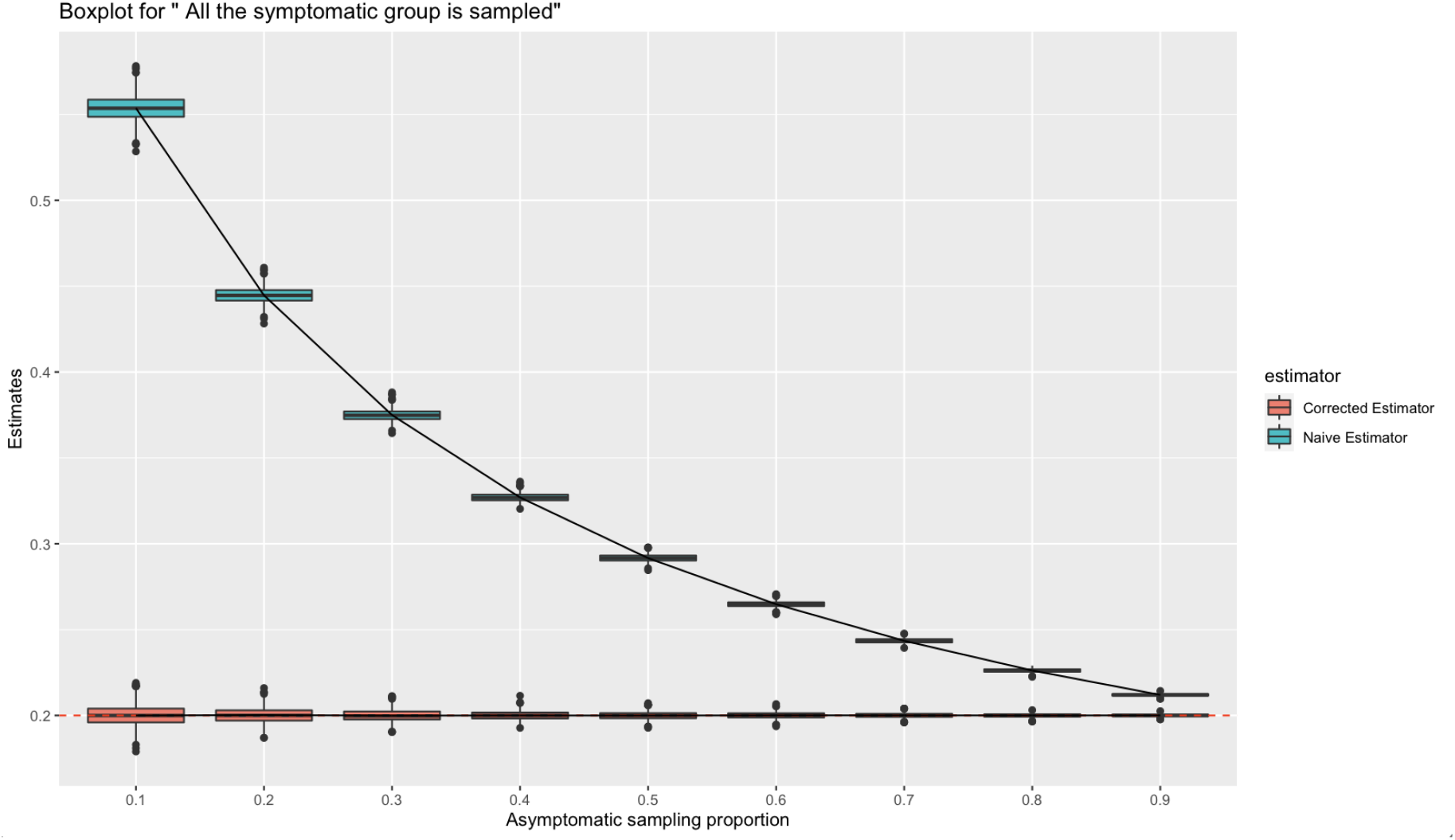
All the symptomatic group is sampled

#### D.2 Not all the symptomatic group is sampled

In this scenario, we assume that only 70% of symptomatic individuals underwent testing. From Figure D2, as the number of asymptomatic individuals in the total testing sample size increases, we observe that the corrected estimator converges to the true value faster than the naïve estimator. Thus we see once again that the testing error correction improves the accuracy of prevalence estimation. By Table D1, it can be seen that our estimated symptomatic rate in population 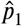 decreases as the number of asymptomatic individuals in the sample increases.

**TABLE D1.** Estimated proportion of symptomatic in the population 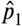 (E(U))

| $\hat{p}_T^{0,*}$ | 0.1 | 0.2 | 0.3 | 0.4 | 0.5 | 0.6 | 0.7 | 0.8 | 0.9 |
| --- | --- | --- | --- | --- | --- | --- | --- | --- | --- |
| $E(U)$ | 0.3881 | 0.3031 | 0.2538 | 0.2221 | 0.1995 | 0.1829 | 0.1701 | 0.1596 | 0.1513 |

**FIGURE D2.**
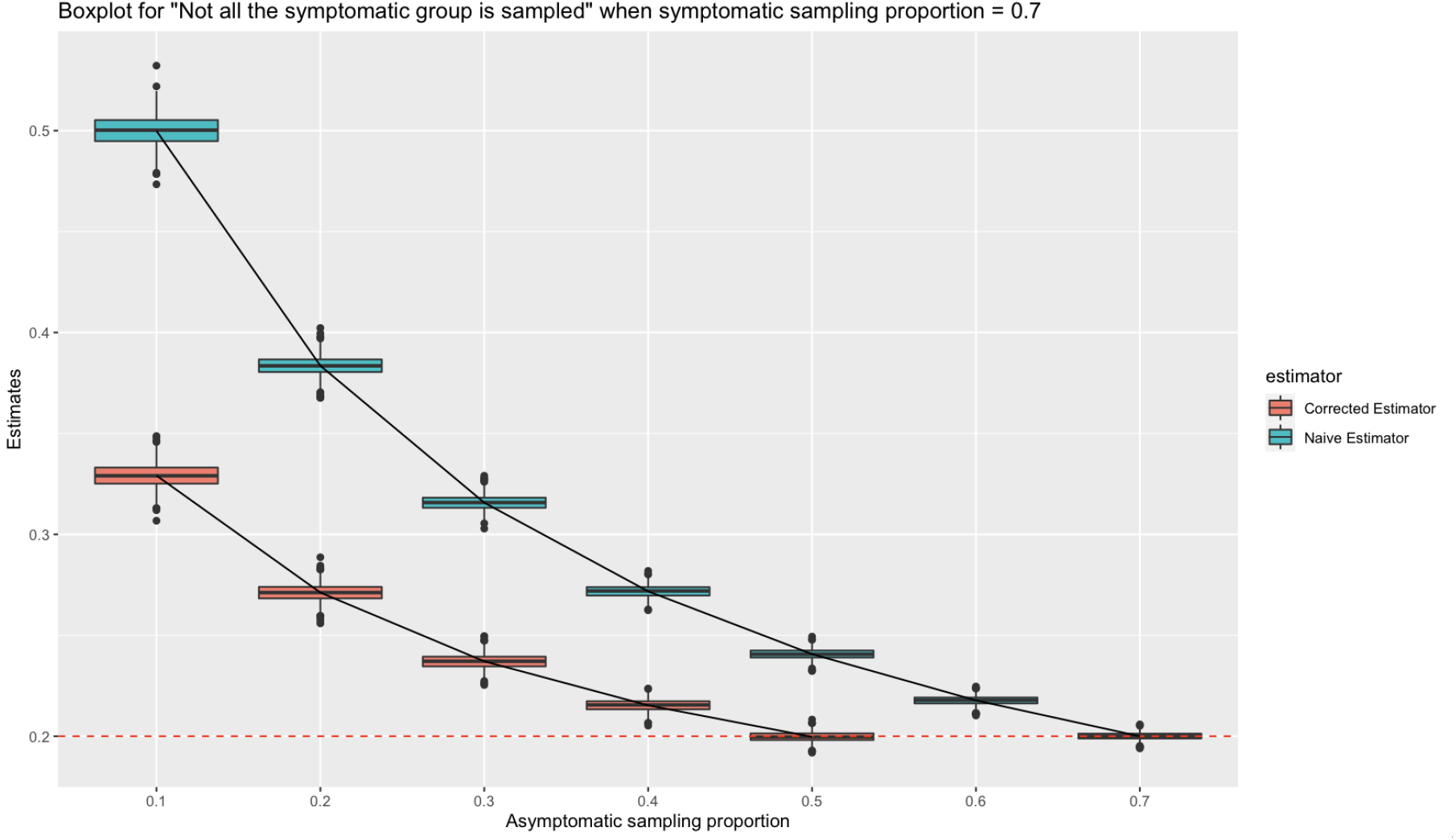
Not all the symptomatic group is sampled

### E CORRECTION OF TESTING ERRORS

In the previous section, we explored the simulation of the correction of sampling error for a population without considering testing errors. In this section, we extend our analysis including testing error for asymptomatic and symptomatic individuals separately. Specifically, we will model the false positive and false negative rates for both groups in testing using normal distributions. The false positive rate for asymptomatic individuals 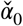 is assumed to follow a normal distribution with mean 0.01 and variance 0.0001, while the false negative rate for asymptomatic individuals 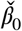 follows a normal distribution with mean 0.2 and variance 0.0001. Similarly, the false positive rate for symptomatic individuals 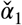 is assumed to follow a normal distribution with mean 0.03 and variance 0.0001, while the false negative rate for symptomatic individuals 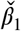 follows a normal distribution with mean 0.02 and variance 0.0001. The real value of the parameters is assumed to be the mean of these distributions. We will consider two scenarios: one in which all symptomatic individuals are sampled, and another in which not all symptomatic individuals are sampled.

#### E.1 All symptomatic group is sampled

In this simulation, we assume that all symptomatic individuals are sampled for the testing group. We will adjust the proportion of asymptomatic individuals getting sampled from 0.1 to 0.9 to observe the effect of testing error correction on prevalence estimation. Based on the description in the previous section, we use the 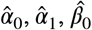 and 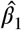 obtained from other study as parameters for testing error correction. Here, we assume that 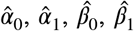 follow a uniform distribution with a mean of the true values *α*_0_ = 0.01, *α*_1_ = 0.03, *β*_0_ = 0.2 and *β*_1_ = 0.02 in the simulation study.

From Figure E3, we expect to see that our corrected estimators are very close to the true value, while the naive estimator is approaching the true value as the proportion of asymptomatic individuals increases. Due to the additional variability introduced by testing error, we observe a larger variance of the corrected estimators compared to Algorithm 1.

#### E.2 Not all the symptomatic group is sampled

We also simulated an scenario where not all symptomatic individuals in the population were sampled for testing, accounting for testing error. Specifically, we assumed that 70% of symptomatic individuals in the population would go for a test.

**FIGURE E3.**
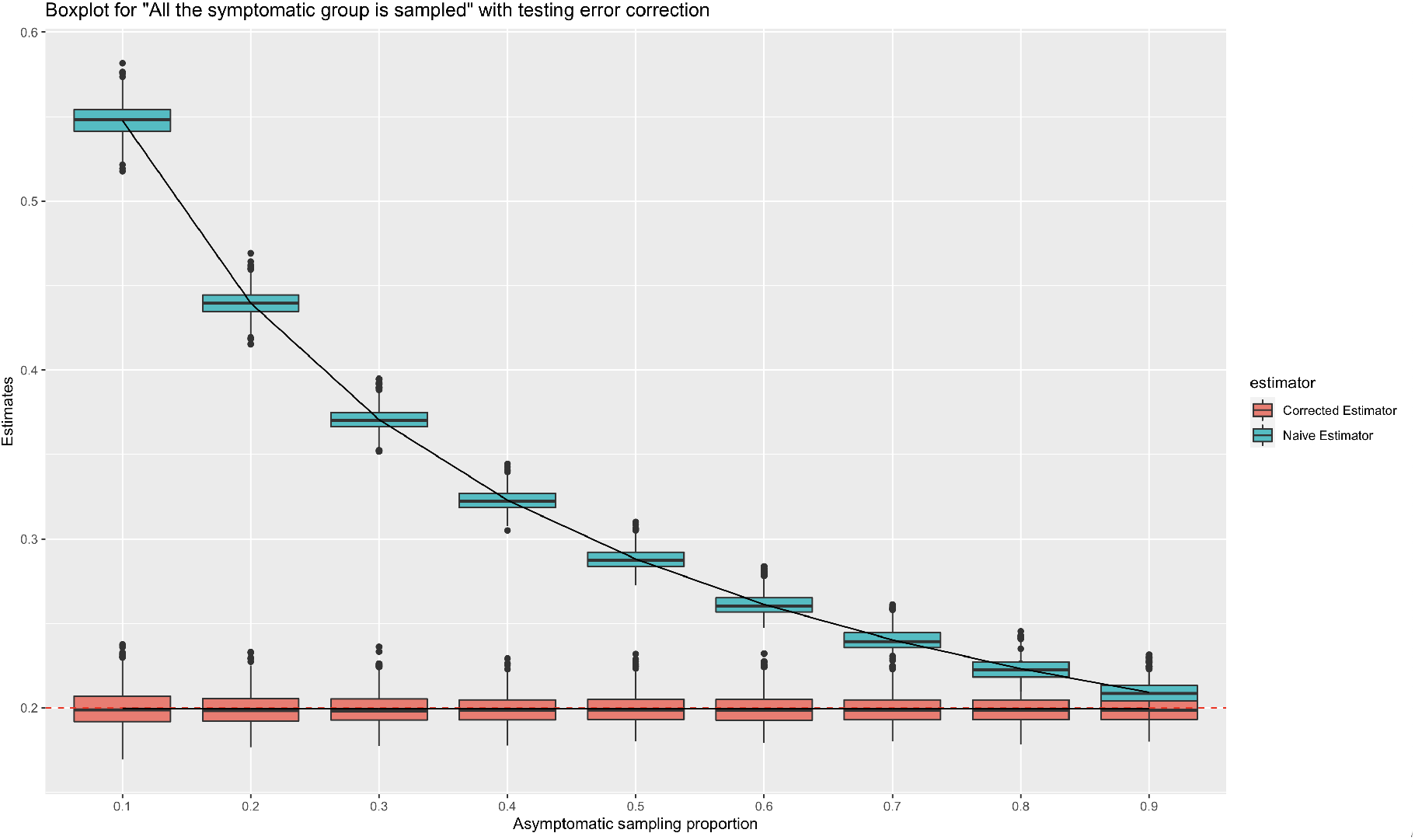
All the symptomatic group is sampled with testing error

**FIGURE E4.**
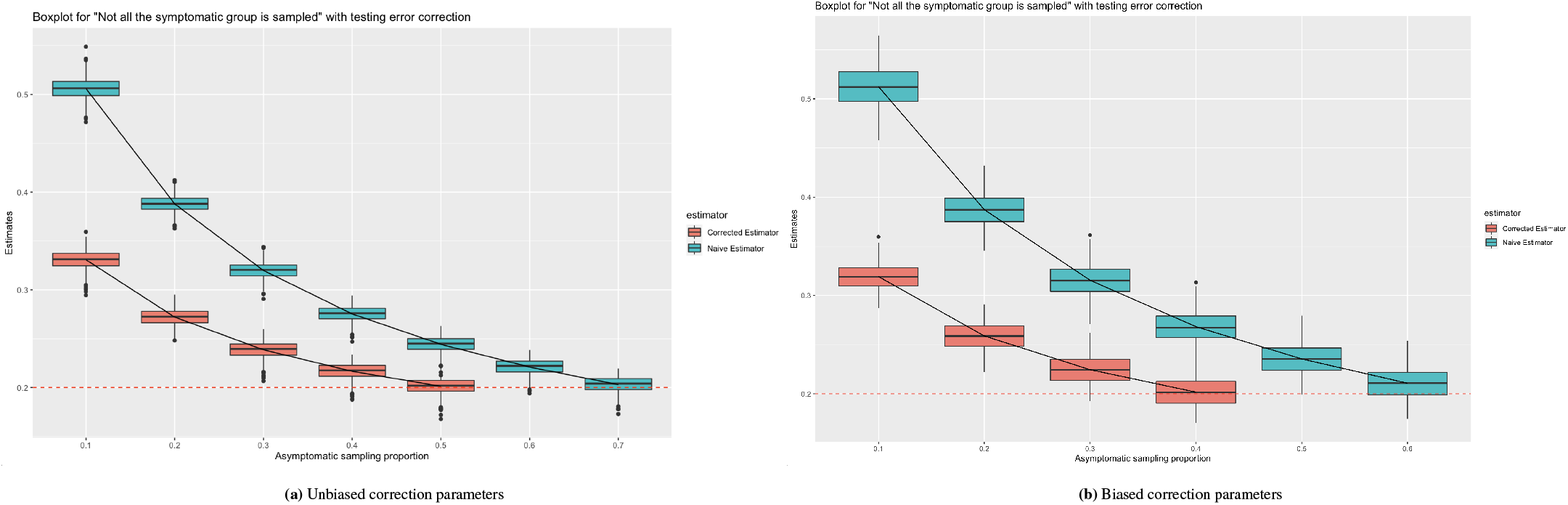
Not all the symptomatic group is sampled with testing error

Here we consider two parameter settings for the correction parameters obtained from other studies: the first one is unbiased, where 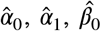 and 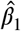 follow a uniform distribution with mean 0.01,0.03,0.2 and 0.02 (the true values), respectively; the second one is biased, where 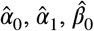 and 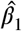 follow a uniform distribution with mean 0.05, 0.1, 0.3, and 0.1, respectively. From the results in Figures E4a and E4b, it can be observed that the estimates from both settings converge to the true values as the proportion of asymptomatic increases in the sample, but the corrected estimate from the second setting has a larger variance.

## Notes

### Competing Interest Statement

The authors have declared no competing interest.

### Funding Statement

DADP, JSR, and CZ acknowledge the support of the Copeland Foundation Award 2022 from the Department of Public Health Sciences at the University of Miami

### Author Declarations

The data used in Section 6 is publicly available at https://github.com/nshomron/covidpred

### Summary of Updates

This new version greatly improves readability and results with respect to the previous version.

